# Trends in homelessness and injection practices among young urban and suburban people who inject drugs: 1997-2017

**DOI:** 10.1101/2021.02.26.21252551

**Authors:** Anna L. Hotton, Mary Ellen Mackesy-Amiti, Basmattee Boodram

**Affiliations:** Department of Medicine, University of Chicago, Chicago, Illinois, USA; Division of Community Health Sciences, School of Public Health, University of Illinois at Chicago, Chicago, Illinois, USA

**Keywords:** people who inject drugs, homelessness, meta-regression, injection risk behavior

## Abstract

**Background:** Among young people who inject drugs (PWID) homelessness is associated with numerous adverse psychosocial and health consequences, including risk of relapse and overdose, psychological distress and suicidality, limited treatment access, and injection practices that increase the risk of HIV and hepatitis C (HCV) transmission. Homeless PWID may also be less likely to access sterile syringes through pharmacies or syringe service programs.

**Methods:** This study applied random-effects meta-regression to examine trends over time in injection risk behaviors and homelessness among young PWID in Chicago and surrounding suburban and rural areas using data from 11 studies collected between 1997 and 2017. In addition, subject-level data were pooled to evaluate the effect of homelessness on risk behaviors across all studies using mixed effects logistic and negative binomial regression with random study effects.

**Results:** There was a significant increase in homelessness among young PWID over time, consistent with the general population trend of increasing youth homelessness. In mixed-effects regression, homelessness was associated with injection risk behaviors (receptive syringe sharing, syringe mediated sharing, equipment sharing) and exchange sex, though we detected no overall changes in risk behavior over time.

**Conclusions:** Increases over time in homelessness among young PWID highlight a need for research to understand factors contributing to youth homelessness to inform HIV/STI, HCV, and overdose prevention and intervention services for this population.

## Background

Homelessness is prevalent among young people who inject drugs (PWID) and has consistently been associated with injection risk behaviors [1-5] and risk for HIV and hepatitis C (HCV) transmission [6-8]. Youth homelessness is also associated with numerous other social adversities and negative health outcomes, including high rates of depression and suicidality, high likelihood of alcohol and substance use problems, poor academic outcomes, sexual risk taking and high rates of sexually transmitted infections (STI), and past and concurrent victimization [9-13]. For young PWID, homelessness is associated with a myriad of adverse effects, including difficulty accessing treatment for substance use [14], risk of relapse [1, 15], blood-borne infections due to syringe and equipment sharing in unsanitary conditions [1, 15], increased risk of overdose [16, 17], vulnerability to violence and sexual exploitation [18, 19], psychological distress and suicidal behavior [20, 21], and general increased morbidity and mortality [22-25]. Homeless PWID may also be less likely to benefit from increased access to sterile syringes at pharmacies in Illinois [26-28] that occurred over the past two decades, either due to lack of resources for purchasing syringes or fear of pharmacy staff or police harassment. PWID from suburban and rural communities who experience homelessness may be at particularly high risk of infectious disease transmission due to less availability of syringe service programs (SSPs) and other harm reduction services compared to urban areas.

Recent data suggest that homelessness has been increasing in the US over the past two decades as a result of economic recessions and rising housing costs [29]. While standard surveys of homelessness do not estimate PWID as a subpopulation, it is possible that increases in homelessness have occurred among PWID as well. Increases in heroin injection among young people as a result of the opioid epidemic may also have resulted in increases in the population of young homeless PWID.

The goal of this study was to examine trends over time in injection risk behaviors and homelessness among young PWID in the Chicago, Illinois and surrounding suburban and rural areas, using data from several federally-funded research studies conducted from 1997 through 2017. Because time was confounded with study, we examined time trends using meta-regression on summary measures. In follow-up analyses, we examined associations between homelessness and risk behaviors across all studies.

## Materials and Methods

### Data sources

De-identified data were obtained from the principal investigators of eleven research studies that collected detailed data on PWID demographics and injection related behaviors over the past twenty years. These included National HIV Behavioral Surveillance (NHBS) PWID cycles 2-4 conducted by the Chicago Department of Public Health (CDPH), the Collaborative Injection Drug User Studies (CIDUS 2 & 3), and local studies conducted by UIC-COIP investigators shown in Table 1. Detailed descriptions of each of the studies in terms of participants, recruitment strategies, and assessment methods have been previously described in the citations listed in Table 1. Briefly, recruitment approaches included either respondent driven sampling (RDS) or convenience sampling via flyers and advertisements posted at the recruitment sites and/or active recruitment by outreach workers. Our analysis was restricted to participants who reported injection drug use. With the exception of NIHU, which focused exclusively on heroin users, eligibility across studies included injection of any substances, though the most commonly reported drug injected was heroin alone or in combination with other drugs (e.g., cocaine, methamphetamine). Harm reduction and drug treatment resources were made available to all participants as part of the study protocols, and many studies involved recruitment directly from SSP sites. CDPH and the principal investigators disclaim responsibility for any analysis, interpretations, or conclusion of the study. This analysis was restricted to young PWID (18-34) because seven of the studies restricted their samples to this age range.

**Table 1.** Data sets included in the metaregression.

| Study | Years | N<br>[ages<br>18- 35] | Measurement time frame |  |  | RDS<br>recruit<br>ment | Percent<br>residence<br>in Chicago |
| --- | --- | --- | --- | --- | --- | --- | --- |
|  |  |  | Homeless | IDU<br>network | Risk<br>behavior |  |  |
| CIDUS 2 [30] | 1997-1999 | 695 | 6 months | NA | 6 months | No | 0.61 |
| CIDUS 3 [31] | 2002-2004 | 775 | 6 months | 3 months | 3 months | No | 0.32 |
| NATHCV [32] | 2002-2005 | 110 | 6 months | NA | 6 months | No | NA |
| NIHU [33] | 2004-2006 | 150 | NA | 30 days | 6 months | No | 0.56 |
| SATHCAP [34] | 2005-2009 | 453 | 12 months | 6 months | 30 days | Yes | 0.62 |
| PSYCH [35] | 2008-2010 | 610 | 6 months | 6 months | 6 months | Yes<sup>a</sup> | 0.28 |
| NHBS IDU2 [36] | 2009 | 143 | 12 months | 30 days | 12 months | Yes | 0.74 |
| NHBS IDU3 [37] | 2012 | 59 | 12 months | 30 days | 12 months | Yes | 0.52 |
| SOCNET [38] | 2012-2013 | 163 | 6 months | 6 months | 6 months | No | 0.80 |
| NHBS IDU4 [39] | 2015 | 263 | 12 months | 30 days | 12 months | Yes | 0.54 |
| MOOD [4] | 2016-2017 | 180 | 6 months | 6 months | 6 months | No | 0.61 |

<sup>a</sup> Partial RDS recruitment; 40% of sample was recruited by outreach
RDS: Respondent driven sampling
CIDUS: Collaborative Injection Drug Users Study (CDC)
NATHCV: Early natural history of HCV infection among injection drug users (NIDA)
NIHU: Non-injected heroin use, HIV, and transitions to injection (NIDA)
SATHCAP: Sexual Acquisition and Transmission of HIV Cooperative Agreement Project (NIDA)
PSYCH: Psychiatric disorders and HIV risk behaviors among young injection drug users (NIDA)
SOCNET: Social networks of young injection drug users (Chicago DCFAR)
MOOD: Emotion dysregulation and risky behavior among people who inject drugs (NIDA)
NHBS: National HIV Behavioral Surveillance (CDC)

### Measures

#### Demographic variables

included sex, race/ethnicity (non-Hispanic White, non-Hispanic Black, Hispanic, and Other, including multiracial categories), and age (categorized as 18-24 vs. 25-34). Subjects identifying as transgender were excluded from the analysis due to insufficient analytic sample size.

#### Harmonization of outcome measures

Data harmonization involved recoding, combining, and collapsing variables as necessary such that constructs were defined and expressed in equivalent scales/units across studies. We did not make any adjustments to account for variation in the time frame for outcome measurement (e.g., past 6 months, past 12 months) across studies (Table 1). Measures from longitudinal studies were summarized across time within individuals such that individuals contributed one observation per study. We created dichotomous indicators for risk behaviors, and converted Likert scale responses on frequency measures to proportions (Never = 0, Rarely = .10, Less than half the time = .25, Half the time = 50, More than half the time = .75, Almost always = .75, Always = 1).

#### Homelessness

was measured as self-perceived homelessness (e.g. have you been homeless, have you considered yourself homeless) within the past 6 or 12 months.

#### Injection networks

Two measures of injection network size were available: 1) total injection network: the number of PWID that the respondent knows and has seen in past 30 days or 6 months (6 studies); and 2) injection sub-network: the number of people the respondent injected drugs with in the past 30 days, 3 months, or 6 months (6 studies). Three studies included both of these measures.

#### Injection-related risk behaviors

Any syringe-mediated drug sharing (SMS) in the past 3, 6, or 12 months was defined as having shot up with a needle after someone else squirted drugs into it from another syringe (i.e., “backloading”), or drugs were mixed, measured, or divided using someone else’s syringe. Receptive syringe sharing (RSS) was defined as injecting in the past 30 days to 12 months with a syringe that someone else had previously used to inject.

Frequency of RSS was estimated as the percent of injections that involved RSS based on either Likert scale responses or the number of RSS injections and frequency of injection. For respondents, we also estimated receptive sharing network size (i.e., the number of people respondent used a syringe after) and distributive sharing network size (i.e., the number of people the respondent gave his/her used syringe to). Equipment sharing was defined as any sharing of cookers, cotton, or rinse water for drug injection in the past 30 days to 12 months. *Exchange sex* was defined as having bought or sold sex for drugs, money, shelter, or other goods in the past 3, 6, or 12 months.

### Statistical analysis

Using the harmonized datasets, frequencies of categorical variables and means and standard errors of continuous variables were computed. Standard errors were calculated based on the Poisson distribution for count variables and the binomial distribution for binary variables. Pooled estimates of means and proportions were computed using the *metan* command in Stata 15.1 with random effects based on the DerSimonian & Laird method and heterogeneity estimated via the Mantel-Haenszel method [40].

For the analysis of time trends, it was necessary to employ meta-regression of summary measures rather than analyzing individual level data due to the association between study and time. Meta-regression is an extension of meta-analysis that can be used to examine whether between-study heterogeneity is related to aggregate level study characteristics (e.g., sociodemographic composition, behavior prevalence, network characteristics) [41]. Using the harmonized datasets, we computed aggregate level summary statistics (means and standard errors) by study for sociodemographic variables, injection behaviors, and network variables. We conducted random-effects meta-regression using aggregate means and standard errors for outcomes of interest. Each study contributes one observation to the analysis, with the number of observations ranging from 5-11 depending on the availability of measures across studies. To examine annual time trends, separate models were fit for each outcome with year as the only independent variable using the *metareg* command in Stata. For ease of interpretation, time was scaled in 10-year intervals so that coefficients represent the average change in the outcome per 10 year period. These analyses were also repeated, stratified by past 6-month homelessness, to determine if trends over time varied according to housing status. Standard errors and confidence intervals for meta-regression coefficients were estimated using the Knapp-Hartung variance estimator [42].

Subject-level data were then pooled to evaluate the effect of homelessness on risk behaviors across all studies. Mixed effects regression models with random study effects were applied, with logistic regression for binary outcomes and negative binomial regression for count outcomes. Analyses were conducted using Stata/SE version 15.1 [43].

## Results

Study characteristics are shown in Table 2. Across studies, participants were mostly non-Hispanic White and male. Approximately one-third were less than 25 years of age and over half resided in Chicago. Homelessness was common (41%) as were injection related risk behaviors with average pooled prevalence of receptive syringe sharing, syringe-mediated drug sharing, and equipment sharing of 38%, 26%, and 65% respectively. Average pooled prevalence of exchange sex was 17%.

**Table 2.** Pooled estimates of young PWID characteristics: 1997-2017.

|  | **N**<br>**studies** | **Pooled mean (95% CI)** | **Range** |
| --- | --- | --- | --- |
| Percent white | 11 | 0.64 (0.58, 0.71) | 0.41 - 0.78 |
| Percent Black | 11 | 0.07 (0.05, 0.10) | 0.02 - 0.30 |
| Percent Hispanic/Latino | 11 | 0.23 (0.19, 0.27) | 0.13 - 0.36 |
| Percent young (<25) | 10* | 0.36 (0.22, 0.50) | 0.11 - 0.63 |
| Percent female | 11 | 0.34 (0.32, 0.36) | 0.27 - 0.42 |
| Percent urban Chicago | 10 | 0.56 (0.45, 0.67) | 0.28 - 0.80 |
| Percent homeless | 10 | 0.41 (0.31, 0.51) | 0.15 - 0.68 |
| Percent receptive syringe sharing | 11 | 0.38 (0.33, 0.43) | 0.27 - 0.49 |
| Percent syringe mediated sharing | 10 | 0.26 (0.23, 0.30) | 0.17 - 0.38 |
| Percent equipment sharing | 9 | 0.65 (0.59, 0.71) | 0.51 - 0.77 |
| Percent exchange sex | 10 | 0.17 (0.14, 0.19) | 0.10 - 0.31 |
| Mean # partners receptive syringe sharing | 7 | 0.73 (0.59, 0.88) | 0.52 - 1.29 |
| Mean # partners distributive syringe sharing | 5 | 1.02 (0.75, 1.30) | 0.69 – 1.95 |
| Mean # partners sharing equipment | 6 | 1.89 (1.36, 2.43) | 1.04 - 2.87 |
| Mean total injection network size | 6 | 13.75 (9.07, 18.44) | 5.08 - 24.48 |
| Mean injection sub-network size | 5 | 4.31 (3.25, 5.36) | 2.82 – 5.46 |

\*PSYCH study excluded: all under 25

In the meta-regression analysis of time trends, we found no significant effects on injection sub-network size (number of people injected with) or any of the injection-related risk behaviors (Table 3). Homelessness among young PWID increased significantly over time (Fig 1, Table 3), with time explaining over 60% of between-study variance in homelessness. Total injection network size (number of people you know who inject drugs) also increased over time; however the effect was not statistically significant. In analyses stratified by homelessness, there were no statistically significant time trends among PWID experiencing homelessness. Among PWID not experiencing homelessness there was a non-statistically significant decline over time in RSS (Coef = −0.10, 95% CI −0.20, 0.004; p=0.057), while among PWID who reported homelessness there was no change in RSS over time (Coef = −0.05, 95% CI −0.18, 0.08; p=0.404). Among PWID not experiencing homeless, the marginal predicted proportion of PWID engaging in RSS was 0.43 in 1997 (95% CI 0.32-0.54) and 0.23 in 2016 (95% CI 0.14-0.33), while among those experiencing homelessness the marginal predicted proportions were 0.53 in 1997 (95% CI 0.39-0.67) and 0.44 in 2016 (95% CI 0.33-0.54).

**Table 3:** Effects of time on risk behavior and homelessness in meta-regression analysis.

| Outcome | N studies | Coefficient<sup>a</sup> | (95% CI) | p-value | Adjusted R-squared<sup>b</sup> |
| --- | --- | --- | --- | --- | --- |
| Percent any receptive syringe sharing | 11 | -0.03 | (-0.14, 0.07) | 0.471 | -3.84% |
| Percent any syringe mediated sharing | 10 | 0.07 | (-0.02, 0.16) | 0.095 | 25.77% |
| Percent any equipment sharing | 9 | -0.01 | (-0.14, 0.11) | 0.825 | -15.43% |
| Percent any exchange sex | 10 | 0.04 | (-0.02, 0.10) | 0.137 | 12.60% |
| Percent homeless | 10 | 0.25 | (0.10, 0.40) | 0.005 | 61.60% |
| Mean # partners receptive syringe sharing | 7 | 0.20 | (-0.42, 0.82) | 0.453 | -6.57% |
| Mean # partners distributive syringe sharing | 5 | 0.56 | (-0.75, 1.87) | 0.266 | 18.34% |
| Mean # partners sharing equipment | 6 | 1.18 | (-0.44, 2.80) | 0.113 | 40.98% |
| Mean total injection network size | 6 | 12.6 | (-13.3, 38.6) | 0.248 | 14.13% |
| Mean injection sub-network size | 5 | 1.36 | (-1.37, 4.09) | 0.212 | 27.81% |

<sup>a</sup> change in outcome per 10-year interval
<sup>b</sup> Adjusted R<sup>2</sup> is the relative reduction in between-study variance explained by adding independent variables (in this case, time) to the model. It is calculated by comparing tau<sup>2</sup> from a model with no variables to tau<sup>2</sup> when variables are added. A negative value indicates that the variables explain less heterogeneity than would be expected by chance.

**Figure 1:**
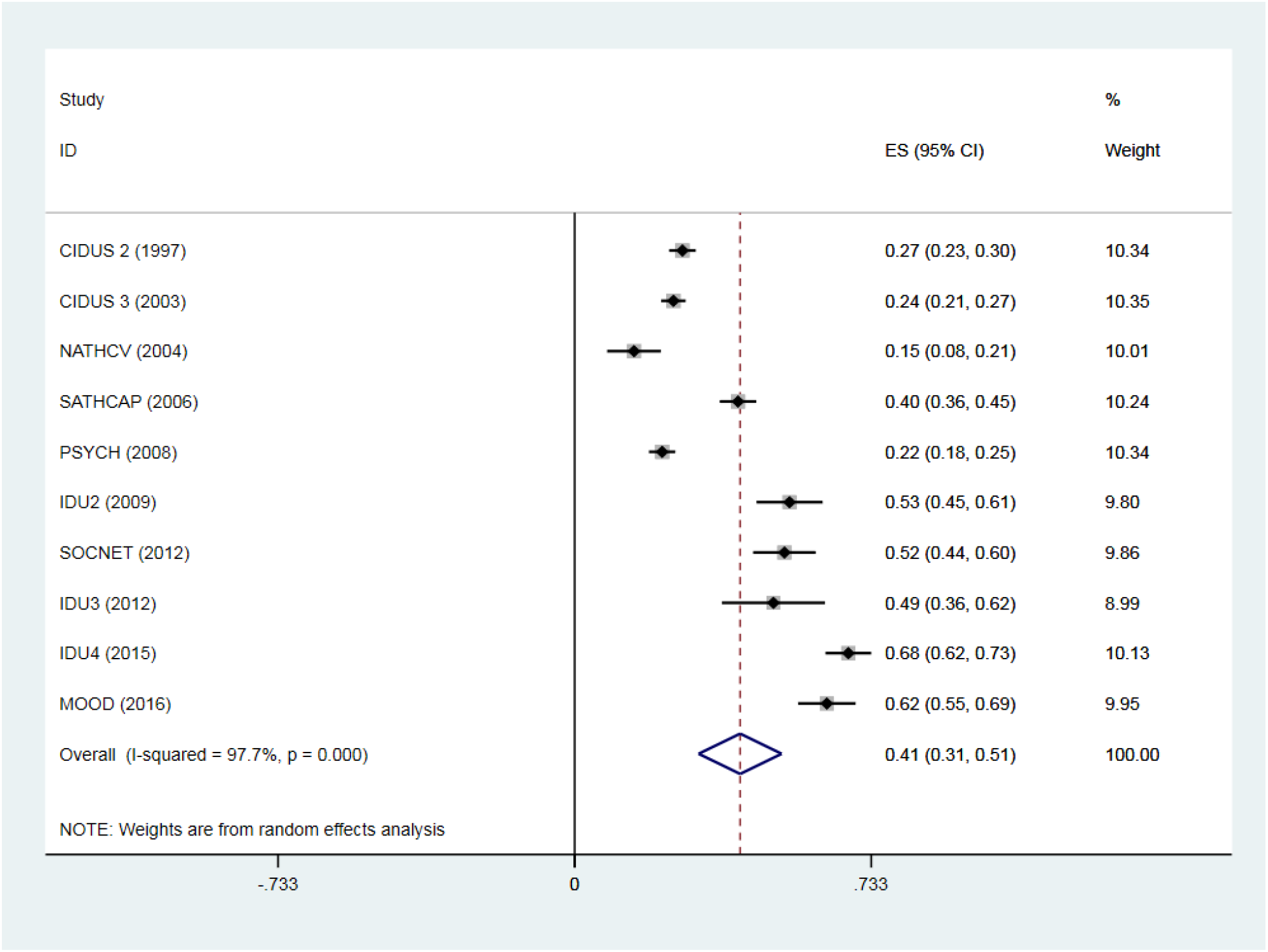
Mean proportion of PWID reporting homelessness by study and year.

We conducted sensitivity analyses on homelessness to assess whether increases in homelessness over time could be partly due to expansion of the reference timeframe (i.e. from 6 months to 12 months) by 1) controlling for the timeframe of assessment of homelessness and 2) restricting the analysis to studies that used past 6 months as the timeframe (n=6 studies).

Findings were similar in all scenarios. With no adjustment (original): Coef = 0.25, 95% CI 0.10, 0.40; p=0.005; with adjustment for timeframe of 6 vs. 12 months: Coef=0.22, 95% CI 0.07-0.37; p=0.012); with restriction to studies with a timeframe of 6 months: Coef=0.21, 95% CI −0.03-0.46; p=0.073.

In subject-level mixed-effects regression of pooled individual-level data with random study intercepts, adjusted for age, race and sex, homelessness was associated with RSS, SMS, equipment sharing, and exchange sex, in addition to total injection network and injection sub-network size and total number of sex partners (Table 4).

**Table 4:** Associations of homelessness with risk behaviors estimated in multivariable mixed effects regression.

|  | Total obs | n<br>groups | aOR or<br>aRR<sup>a</sup> | (95% CI) | p-value |
| --- | --- | --- | --- | --- | --- |
| *Binary outcomes* |  |  |  |  |  |
| Receptive syringe sharing | 3429 | 10 | 1.93 | (1.67-2.23) | <0.001 |
| Syringe mediated sharing | 2699 | 9 | 2.18 | (1.81-2.63) | <0.001 |
| Equipment sharing | 3326 | 9 | 1.71 | (1.43-2.05) | <0.001 |
| Exchange sex | 3321 | 9 | 2.45 | (2.17-2.76) | <0.001 |
| *Count outcomes* |  |  |  |  |  |
| Total RSS partners | 2459 | 7 | 2.15 | (1.48-3.13) | <0.001 |
| Total DSS partners | 2261 | 5 | 2.24 | (1.60-3.14) | <0.001 |
| Total injection network size | 1665 | 6 | 1.55 | (1.38-1.73) | <0.001 |
| Injection sub-network size | 2144 | 5 | 1.48 | (1.25-1.76) | <0.001 |
| Total sex partners | 3288 | 9 | 1.41 | (1.18-1.69) | <0.001 |
| Total exchange sex partners | 1609 | 5 | 3.32 | (2.57-4.28) | <0.001 |

a Adjusted for age, race, and sex. Estimates generated from mixed effects logistic or negative binomial regression with random study intercepts.

## Discussion

This analysis showed a significant increase in homelessness among young PWID over time, consistent with the general population trend of increasing youth homelessness[29]. Although homelessness was associated with risk behavior across time, we detected no overall change in the level of risk behavior over time. A partial explanation may lie in the expansion of interventions that increased access to sterile syringes and harm reduction education in Chicago and surrounding areas, which may have led to decreased risk behavior, thereby offsetting the increased homelessness among PWID. Stratified samples by homelessness revealed small decreases in RSS over time, but only among young PWID not experiencing homelessness. Although the observed effects were not statistically significant, it suggests that gains in harm reduction may have been countered by worsening social conditions over time. Another potential driving factor may be increasing proportions of suburban PWID over time, who report having less access to SSPs. Increasing homelessness may be explained partly by differences in sampling, increasing outreach efforts to reach homeless PWID, or increasing representation of socially and economically marginalized populations in study samples over time. Further research is needed to understand causes of the observed time trends in homelessness. Furthermore, explicit homelessness is one outcome along a continuum of housing instability that could include for example couch surfing and doubling up. PWID may experience varying degrees of housing instability, and changes in housing status over time. Future studies could expand definitions of homelessness to gain more nuanced information about how different types of housing instability impact injection risk.

Our findings are consistent with other studies linking homelessness with injection risk behavior, an association that appears to be consistent across age groups, geographic locations, and time periods [1, 8, 44, 45]. Developmentally tailored interventions to increase access to harm reduction services and address the causes of homelessness among young PWID are necessary for prevention of HIV/HCV, overdose, and other negative health and social consequences. Increasing access to mental health services for young PWID is also a priority given the high rates of mental illness among young PWID[4, 35], particularly those experiencing homelessness [46].

### Limitations

An important limitation of this study is the small number of samples included in the meta-regression that limited our ability to detect changes in PWID behavior over time. Findings may not be generalizable to PWID in other geographic locations or settings. Participants were also predominantly non-Hispanic white and samples contained small proportions of Black and Hispanic PWID. Thus we did not have sufficient power for comparison of trends over time by race/ethnicity or for subgroup analyses. Understanding risk associated with homelessness and other stressors unique to PWID from Black and Hispanic communities remains a priority for future research. Because many of the included studies recruited participants from an urban SSP, participants likely had more frequent contact with harm reduction services than would be expected in other PWID, which may partly explain why increases in injection risk were not observed despite increases in homelessness. While measures were harmonized to be as equivalent as possible across studies, differences across studies in time frame of measurement and definitions may have contributed to between study heterogeneity in prevalence of homelessness and injection behaviors and network size. For example, variation in the time frame for assessment of the size of social and injection networks is problematic since networks are often dynamic and may change over short periods due to factors such as incarceration, drug treatment, or mortality. Furthermore, if the timeframe for homelessness increased over time (e.g., from past 6 months to past 12 months), apparent increases in homelessness could have been due to expansion of the reference timeframe. The magnitude of the time trend in homelessness was slightly attenuated with adjustment for the assessment timeframe, but the result remained statistically significant. Restriction to samples with an assessment timeframe of 6 months also yielded similar results. Thus, these findings suggest that the increasing trend in homelessness was not explained solely by differences in the timeframe of measurement.

## Conclusions

Increases over time in homelessness among young PWID are concerning and highlight the need to understand the correlates of youth homelessness among general and at-risk populations to inform appropriate targeting of HIV/HCV/STI/overdose prevention and intervention services. Age appropriate interventions for young PWID are critical to prevent them from falling into chronic homelessness.

## Data Availability

Data are available from the author upon request

